# Machine learning and natural language processing for the early detection of potential mental disorders among school-age children: a prospective birth cohort study

**DOI:** 10.1101/2025.09.10.25335509

**Authors:** Shanquan Chen, Ting Dang, Mengjie Qian, Huizhi Liang, Diribsa Tsegaye Bedada, Quinette Abegail Louw, Anna Moore, Rudolf N. Cardinal, Tamsin J. Ford

**Affiliations:** International Centre for Evidence in Disability, London School of Hygiene & Tropical Medicine, London, United Kingdom, WC1E 7HT; School of Computing and Information Systems, University of Melbourne, Melbourne, Australia, VIC 3010; Department of Engineering, University of Cambridge, Cambridge, United Kingdom, CB2 1PZ; School of Computing, Newcastle University, United Kingdom; Department of Health and Rehabilitation Sciences, Faculty of Medicine and Health Sciences, Stellenbosch University, Cape Town, South Africa; Department of Statistics and Actuarial Science, University of Waterloo,Canada; Department of Health and Rehabilitation Sciences, Faculty of Medicine and Health Sciences, Stellenbosch University, Cape Town, South Africa, 7505; Department of Psychiatry, University of Cambridge, Cambridge, United Kingdom, CB2 0SZ; Cambridgeshire and Peterborough NHS Foundation Trust, United Kingdom, CB21 5EF

**Author notes:** **Corresponding author: Shanquan Chen, assistant professor, PhD**, International Centre for Evidence in Disability, London School of Hygiene & Tropical Medicine, London, United Kingdom, WC1E 7HT.

**Keywords:** Machine learning, Natural language processing, Mental health screening, Children

## Abstract

**Background:** Early detection of childhood mental health disorders remains challenging due to gaps in current screening approaches that lack sensitivity to subtle psychological indicators and rely heavily on observable behaviors. We investigated whether integrating machine learning with natural language processing of children’s written expressions could enhance early detection of potential mental disorders among school-age children.

**Methods:** This prospective birth cohort study used National Child Development Study (NCDS) data, analyzing 8,981 children born in 1958 in the United Kingdom. Mental health outcomes were assessed using the Bristol Social Adjustment Guide (BSAG) and Rutter A Scale at age 11, with cases defined by scores above 95th and 90th percentiles. Predictive models combined traditional risk factors with natural language features extracted from children’s essays describing their imagined future at age 25. We developed eight machine learning models using various predictor combinations, evaluating performance through receiver operating characteristic (ROC) values.

**Results:** Using BSAG 95th percentile threshold, models combining top five selected variables with essay features achieved significantly higher predictive capability (ROC:0.77, 95%CI:0.71-0.83) compared to models using all variables (ROC:0.70, 95%CI:0.63-0.76) or essay features alone (ROC:0.67, 95%CI:0.60-0.74). At 90th percentile threshold, this integrated approach showed similar improvement (ROC:0.81, 95%CI:0.78-0.85). Key predictors included gestational length, maternal parity, parental age, residential characteristics, parental engagement metrics, and children’s BMI. Sensitivity analyses using Rutter A Scale confirmed these findings.

**Conclusion:** Combining machine learning with natural language processing of children’s future-oriented essays offers a promising approach for early detection of childhood mental health disorders. This integrated screening method could facilitate more timely intervention, though validation in contemporary populations is needed before clinical implementation.

## Introduction

Mental health disorders in childhood represent a critical public health concern with far-reaching implications for individual development and societal well-being. Approximately 50% of mental health conditions manifest before age 14, significantly impacting educational attainment, social relationships, and long-term occupational outcomes(1). These early-onset conditions often predict increased risk for physical health complications, substance use disorders, and reduced quality of life in adulthood(2-4), underlining the paramount importance of early identification and intervention.

Despite the well-documented significance of early mental health support, substantial underdiagnosis exists among school-age children(5, 6). While 10-20% of children worldwide experience clinically significant mental health problems(5), only about one-third receive appropriate diagnosis and treatment(6). This diagnostic gap stems from limited access to mental health professionals, stigma surrounding mental health, and inherent challenges in identifying mental health concerns in young populations(5-8). Young children typically lack the emotional vocabulary and self-awareness to articulate psychological distress, instead expressing difficulties through behavioral changes, physical complaints, or academic struggles. Additionally, they depend on caregivers to recognize these signs and initiate appropriate assessment.

Traditional diagnostic approaches such as clinical interviews and standardized assessments rely heavily on observable behavioral symptoms or parent/teacher reports, potentially missing subtle early indicators of psychological distress. Conventional screening methods are also resource-intensive and time-consuming, making widespread implementation challenging. The effectiveness of observational screening is highly dependent on healthcare worker training and confidence(9), while screening tools developed in Western settings may not adequately reflect mental health manifestations in diverse cultural contexts(10, 11).

Machine learning and natural language processing (NLP) technologies offer promising avenues for enhancing early detection of childhood mental health concerns by analyzing complex patterns across multiple data sources(12-16). However, current approaches face two distinct challenges. First, automatic screening tools using commonly recognized risk factors remain limited in accuracy and scalability(17). Second, there is critical need to explore children’s inner psychological experiences that are inaccessible through conventional assessment methods. Research has established connections between language development and mental health in children(18, 19). Language patterns—including vocabulary usage, emotional tone, and narrative coherence—can reveal underlying psychological states when children cannot directly express their emotions(18, 19). Children’s inner world often remains implicit and easily overlooked from an adult perspective, with traditional observable indicators potentially missing subtle psychological manifestations. Natural language processing of children’s written expressions offers a unique window into their psychological well-being, potentially capturing indicators of emotional and mental states missed through conventional observational methods.

The present study aims to evaluate the effectiveness of integrated machine learning and natural language processing approaches for early detection of potential mental disorders among school-age children. While recent research has applied NLP to NCDS data for predicting outcomes such as reading comprehension(20), application specifically for childhood mental health prediction remains underdeveloped. We address three key research questions: (1) To what extent can machine learning algorithms predict childhood mental health outcomes using traditional risk factors? (2) What is the predictive value of natural language processing analysis of children’s written narratives? (3) Does integrating machine learning with natural language processing features enhance predictive accuracy compared to either approach independently? Through addressing these questions, we aim to develop an approach facilitating earlier identification of children at risk for mental health disorders, ultimately enabling more timely intervention and improved outcomes.

## Methods

### Study design and participants

This prospective birth cohort study utilized data from the National Child Development Study (NCDS), which follows 18,558 individuals born during one week (March 3-9) in 1958 across England, Wales, and Scotland(21). The NCDS was selected for its unique collection of children’s written essays at age 11, offering invaluable linguistic data unavailable in newer cohorts. We focused on data from three assessment periods: birth, age 7, and age 11.

At birth, detailed information was collected regarding prenatal conditions, birth characteristics, and parental characteristics. Subsequent waves at ages 7 and 11 captured physical, educational, and social development, economic circumstances, family employment, family life, health behaviours, and wellbeing. Of particular importance was an essay-writing task administered at age 11, where participants responded to: “Imagine that you are now 25 years old. Write about the life you are leading, your interests, your home life, and your work at the age of 25.” This task was completed under standardized conditions in school classrooms with 30 minutes allocated.

We included participants who completed the age-11 essay task. Participants were excluded if they had more than 30% missing values across variables of interest or if their essays contained fewer than 50 words. The final analytical sample and participant flow are presented in **Figure 1**. A comprehensive description of NCDS methodology has been published previously by Power and Elliott(21).

### Outcomes and measures

#### School-children mental health

The primary outcome was childhood mental health, assessed using two validated instruments: the Bristol Social Adjustment Guide (BSAG) and the Rutter A Scale, with the latter serving as sensitivity analysis.

The BSAG, administered by teachers at ages 7 and 11, evaluates children’s social adjustment and behavior in educational settings. This instrument encompasses 12 behavioral domains: unforthcomingness, withdrawal, depression, anxiety for acceptance by adults, hostility towards adults, writing off of adults and adult standards, anxiety for acceptance by children, hostility towards children, restlessness, inconsequential behaviour, and miscellaneous symptoms (both general and nervous)(22, 23). Scoring sums coded items across domains, with higher scores indicating more pronounced mental and behavioral difficulties.

The Rutter A Scale, completed by mothers at ages 7 and 11, assesses childhood mental health problems using a modified version consisting of 14 items on a three-point Likert scale (0 = ‘Does not apply’, 1 = ‘Applies somewhat’, 2 = ‘Certainly applies’)(24). The scale evaluates emotional problems, peer problems, behavioral problems, hyperactivity, and prosocial behaviour, including items on fidgeting, destructive behavior, fighting, worry, social preferences, irritability, emotional state, physical mannerisms, thumb-sucking, nail-biting, disobedience, attention difficulties, adaptation to new situations, and peer victimization(24). The instrument demonstrates good psychometric properties, with reported inter-rater reliability (r = 0.64) and test-retest reliability (r = 0.74)(25).

Potential mental disorders at age 11 were operationalized using threshold-based approaches. Cases were classified using both 95th and 90th percentile thresholds of total aggregate BSAG scores, with parallel analyses conducted using Rutter A Scale scores at the same thresholds to assess model robustness.

### Potential predictors

Our selection of potential predictors was informed by previous research and encompassed three primary domains: socioeconomic position, adverse childhood experiences, and environmental factors.

#### Socioeconomic position

We analyzed multiple harmonized indicators of socioeconomic position from the NCDS. Paternal occupation and employment status were assessed at birth, age 7, and age 11, using the 1990

Registrar General’s Social Class system(26, 27). Occupations were categorized as manual or non-manual, and employment as employed or unemployed(26, 27). Maternal and paternal education was measured by completion of post-compulsory education, which distinguishing between those who remained in education beyond the compulsory period and those who left at the minimum permissible age (26, 27).

Housing tenure at ages 7 and 11 distinguished between owned/mortgaged properties and other arrangements(26, 27). Additional indicators included bedroom count at ages seven and eleven, housing difficulties, financial hardships, and free school meals eligibility. These measures provided comprehensive assessment of socioeconomic circumstances throughout childhood(28, 29).

#### Adverse childhood experiences (ACEs)

Adverse childhood experiences (ACEs) were operationalized based on established definitions from the National Child Development Study (NCDS) research (30, 31). These experiences are defined as traumatic and stressful psychosocial conditions within the familial environment (30, 31). Such conditions typically share three key characteristics: they tend to co-occur, persist over time, and remain outside the child’s control. Survey data were collected at two time points, ages seven and eleven years, capturing seven distinct dimensions of childhood adversity. These dimensions encompassed parental separation or divorce, substance misuse by parents, presence of family conflict, parental death, mental health problems among parents, physical neglect experienced from parents, and parental involvement in criminal activities. Each dimension was assessed as a binary variable, creating a comprehensive framework for evaluating early-life adversity. The methodology for measuring these adversities has been extensively documented in previous publications, with detailed protocols available for references(30, 31).

#### Environmental factors

The analysis incorporated a set of established environmental and psychosocial risk factors associated with mental health outcomes(28, 29). Perinatal factors included infant health conditions at birth (such as haemolytic and respiratory diseases), sex, body mass index z-scores (Zbmi, measured at birth, age seven, and eleven, calculated according to WHO age- and sex-standardized reference values(32)), and maternal health conditions during pregnancy including hypertension and diabetes. Maternal characteristics encompassed age at birth, parity, length of gestational period, marital status at birth, and smoking behavior during pregnancy.

Parental engagement was assessed through multiple dimensions across two time points (ages seven and eleven). For mothers, these dimensions included interest in the child’s education, engagement in outdoor activities such as walking or park visits, and participation in reading activities. Similarly, father’s involvement was evaluated through educational interest, participation in childcare, engagement in outdoor activities, and reading behaviors.

Detailed measurement protocols for all environmental factors are available in previous publications(28, 29).

Demographic variables such as age and race were not included as covariates due to the homogeneous nature of the cohort: all participants were born in March 1958, and the sample was predominantly white (98.7%).

### Missing Data

Cases with substantial missing data, those lacking information for 30% or more variables of interest, were excluded from the analysis. For the remaining cases, multiple imputation with chained equations were employed to generate 25 imputed datasets using all variables of interest as predictors in the imputation model. The multiple imputation approach was selected to address potential bias from missing data patterns while preserving the statistical relationships among variables and maintaining analytical power.

### Statistics and Modeling

Categorical variables were reported as numbers (percentages) and continuous variables as means (standard deviations).

Our modeling approach used mental health outcomes at age 11 (BSAG and Rutter A Scale scores) as the dependent variables, while incorporating predictor variables collected across multiple time points—specifically at birth, age 7, and age 11.

The cohort data, comprising outcomes, potential predictors, and essays, was randomly partitioned into training (80%) and test (20%) datasets. To address class imbalance in the training dataset, given the relatively small number of children with potential mental disorders, we employed the synthetic minority oversampling technique (SMOTE) to generate synthetic samples for the minority class(33).

To maximize practice utility, potential predictors were maintained in their original coded form rather than being aggregated (such as summing the seven ACEs) or transformed through dimensional reduction techniques (such as principal component analysis for socioeconomic position). To avoid overfitting, feature selection was performed using recursive feature elimination (RFE) with the caretFuncs algorithm from the ‘caret’ package(34). This model-agnostic approach was chosen to ensure selected variables would be compatible across multiple machine learning algorithms. The importance scores of selected predictors were derived using this RFE process, which ranks variables based on their contribution to model performance when iteratively removed. The RFE process was optimized through 5-fold cross-validation in the training dataset, with the optimal number of features determined by the maximum ROC value in the test dataset.

For natural language processing, the children’s essays underwent standardized preprocessing steps including lowercase conversion, punctuation removal, stop word filtering, and tokenization to normalize the text data. Then, essays were embedded using the “all-mpnet-base-v2” model, which is based on a pretrained transformer language model architecture developed by Microsoft(35). This model was chosen for its superior performance in capturing semantic relationships and contextual information in text data(35). Unlike traditional word-level embeddings, this sentence-transformer model generates contextualized representations that capture the nuanced meaning of complete sentences and paragraphs. The resulting high-dimensional embeddings (exceeding 700 dimensions) were reduced using uniform manifold approximation and projection (UMAP), selected for its ability to preserve both local and global structure while maintaining computational efficiency(36). We explored reduced dimensionality ranging from 5 to 25 components, with the dimension-reduced essays also evaluated using caretFuncs from the ‘caret’ package. This process underwent 5-fold cross-validation in the training dataset, with optimal UMAP components determined by maximizing ROC values in the test dataset.

Eight prediction models were developed to ensure our findings were not model-specific and to identify the most robust approach for mental health prediction: linear approaches (linear discriminant analysis using the “sda” method and logistic regression using “glm” with the binomial family), non-linear algorithms (classification and regression trees using “xgbTree”, k-nearest neighbours using “knn”, neural networks using “nnet”, and naive Bayes using “naive_bayes”), and advanced ensemble methods (support vector machines using “svmRadial” and random forest using “rf”). All models were implemented using the ‘caret’ R package and underwent 5-fold cross-validation during training. The models were trained using six distinct predictor combinations: all selected features; all UMAP components; and four incremental combinations pairing UMAP components with top selected features (top 5, 10, 15, and 20 features where applicable).

Trained models’ performance was assessed exclusively on the test dataset using receiver operating characteristic (ROC) values. Other metrics including sensitivity, specificity, accuracy, AUC (area under the ROC curve), F-score, kappa statistic, precision, and recall were also provided.

These modeling processes were replicated for each mental health outcome measure. All analyses were conducted using R version 4.3, with essay embedding performed in Python.

## Results

In this study, 8,981 children (49.3% female) were included. **Table 1** presents the basic characteristics of included participants across three key domains: socioeconomic position, adverse childhood experiences, and environmental factors. Socioeconomic indicators revealed a predominance of manual occupations among fathers (71.9% at birth, declining to 64.5% at age 11), with approximately one-third of parents pursuing post-compulsory education. Housing stability improved moderately through childhood, as evidenced by increased home ownership from 42.1% to 46.0% between ages 7 and 11. Notable adverse childhood experiences included family conflict and parental physical neglect (both 6.3%), alongside parental mental health challenges (6.2%). Environmental factors demonstrated high parental engagement, with sustained maternal interest in children’s education (85.7% at age 7, 86.2% at age 11) and widespread participation in outdoor activities. Most mothers were in stable unions (96.8%), though 23.3% experienced pregnancy-related health conditions. Birth weight and BMI distributions predominantly fell within normal ranges across all assessment points.

Mental health outcomes at age 11, measured via the Bristol Social Adjustment Guide and Rutter A Scale, yielded mean scores of 15.81 (SD=17.01) and 6.56 (SD=3.65), respectively (**Table 1**).

Figure 2 illustrates the relative predictive importance of variables for childhood mental health outcomes, stratified by two BSAG score thresholds (95th and 90th percentiles). For the more stringent 95th percentile threshold (**Panel A**), gestational period emerged as the strongest predictor, followed by parental age at birth and residential characteristics. The analysis revealed that environmental factors, particularly those related to early-life conditions, demonstrated higher predictive importance than socioeconomic indicators. Parental engagement metrics, including father’s interest in children’s education and maternal smoking during pregnancy, ranked among the top ten predictive factors. The analysis at the broader threshold (**Panel B**, 90th percentile) corroborated these findings, with consistent prominence of the identified key predictive factors.

**Supplementary Table 1** presents a comparison between included and excluded children. Children excluded from the analysis (those with more than 30% missing values or essays containing fewer than 50 words) exhibited significantly poorer mental health than their included counterparts (p values<0.01).

Figure 3 and **Table 2** demonstrate the comparative performance metrics of machine learning models in detecting childhood mental health disorders. The analysis, stratified by BSAG score thresholds at the 95th and 90th percentiles, revealed significant improvements through feature integration. At the more stringent 95th percentile threshold, the combination of top 5 selected variables with essay features yielded substantial enhancement in predictive capability (ROC: 0.77, 95% CI: 0.71-0.83), markedly outperforming models using either all selected variables (ROC: 0.70, 95% CI: 0.63-0.76) or essay features alone (ROC: 0.67, 95% CI: 0.60-0.74). Model performance was further refined through incremental feature integration. The incorporation of top 10 selected variables with essay features preserved the enhanced predictive capacity while improving specificity metrics (0.78, 95% CI: 0.76-0.80). Additional optimization was achieved through the integration of top 15 and 20 selected variables, ultimately reaching peak ROC values of 0.81 (95% CI: 0.76-0.86). Parallel analyses at the 90th percentile threshold exhibited analogous patterns of improvement. The integration of top 5 selected variables with essay features demonstrated marked enhancement in model performance (ROC: 0.81, 95% CI: 0.78-0.85) over single-feature approaches. This performance was slightly elevated through the incorporation of top 10 selected variables (ROC: 0.82, 95% CI: 0.79-0.85), with subsequent feature additions maintaining superior predictive metrics while optimizing sensitivity and specificity parameters.

The sensitivity analysis using Rutter A Scale scores corroborated our primary findings. The analysis demonstrated similar patterns in feature selection importance (**Supplementary Figure 1**) and progressive enhancement in machine learning model performance through the combined use of multiple feature types (**Supplementary Figure 2** and **Supplementary Table 2**), specifically, the additive combination of top 5 selected variables with essay features also yielded substantial improvements in predictive capability.

## Discussion

In this prospective birth cohort study leveraging data from the National Child Development Study (NCDS), we examined the predictive capability of machine learning and natural language processing approaches for early detection of potential mental disorders among school-age children. Our findings demonstrated that the integration of traditional risk factors with natural language features derived from childhood essays significantly enhanced predictive performance, with the most notable improvement observed in the parsimonious combination of the top 5 selected variables and essay features. This integrated approach outperformed models using either all selected variables or essay features alone, even though subsequent feature integration achieved incremental improvements. Environmental factors, particularly gestational period, maternal parity, parental age at birth, and early-life socioeconomic status (e.g. residential bedroom count), emerged as the strongest predictors. Parental engagement metrics, including father’s interest in children’s education and maternal smoking during pregnancy, as well as children’s zBMI almost ranked among the top ten predictive factors. The robustness of these findings was further validated through sensitivity analyses using Rutter A Scale scores, underscoring the potential utility of combining targeted traditional risk factors with natural language processing for efficient early mental health screening in pediatric populations.

The integration of traditional risk factors with natural language features derived from childhood essays demonstrated superior predictive performance for early detection of mental health concerns, with optimal results achieved through a parsimonious combination of five key variables and essay features. This finding aligns with previous research, which demonstrated the value of multimodal approaches in psychiatric assessment(13, 37, 38). However, while prior studies typically emphasized the need for comprehensive data collection across multiple domains, our results suggest that a more streamlined approach may achieve comparable predictive accuracy. This apparent divergence might be attributed to our novel application of advanced natural language processing techniques, which potentially capture more nuanced psychological indicators than traditional assessment methods. For example, our approach may detect subtle linguistic patterns such as metaphor usage, emotional tone shifts, self-referential language, and future orientation that could reflect underlying psychological states not readily observable through conventional behavioral assessments or parental reports. The superior performance of our integrated approach compared to single-modality models corroborates previous meta-analytic findings, which identified significant advantages in combining behavioral and linguistic markers for clinical assessment(39-41).

Our analysis revealed the predominant predictive power of environmental factors, particularly perinatal and early-life conditions, in determining childhood mental health outcomes. The emergence of gestational period as a primary predictor corresponds with previous longitudinal studies, which established robust associations between prenatal development and subsequent psychological outcomes(42-44). The significant predictive value of parental characteristics (e.g. maternal parity and parental age) and the parental engagement metrics (e.g. paternal involvement) extends previous findings of parental influences on mental health or psychological development of children(45-47). However, while earlier research emphasized socioeconomic indicators as primary predictors(48, 49), our results suggest that environmental factors may have greater predictive utility. This discrepancy might be attributed to our study’s assessment of early-life conditions and the inclusion of subtle indicators such as residential characteristics.

The implications of these findings extend across potential clinical practice, public health policy, and future research directions. First, it is important to note that our study assessed school functioning and behavioral adjustment rather than formal clinical diagnoses of mental health disorders. The BSAG and Rutter A Scale measure behavioral and social adjustment problems that, while strongly correlated with mental health conditions, represent functional outcomes rather than diagnostic classifications. This distinction is critical when interpreting our findings within the broader context of mental health screening. Our integrated approach demonstrated strong predictive performance, it should be viewed as a complement to, rather than a replacement for, comprehensive clinical evaluation. While both BSAG and Rutter scales were collected at the same time as the essays in our study, it is worth noting that these traditional scales usually depend on teachers, parents, or caregivers and often require specific training for proper utilization. In contrast, the essay-based approach demonstrated in our study offers potential for wider application with less dependency on observer training or subjective interpretation. The model’s primary utility lies in its potential to efficiently identify children who might benefit from more detailed clinical assessment, thereby enabling earlier intervention for those at greatest risk. The demonstrated efficacy of integrating machine learning with natural language processing suggests a potential approach to early mental health screening in pediatric populations.

If established to be useful, this integrated methodology could facilitate more efficient resource allocation in mental health services by enabling targeted interventions for high-risk children, particularly in resource-constrained settings where comprehensive clinical assessments may be limited. Furthermore, the identification of key environmental predictors, especially those related to perinatal and early-life conditions, underscores the importance of preventive interventions during critical developmental periods. From a practical implementation perspective, it is noteworthy that several of our strongest predictors – such as gestational period, residential bedroom count, and parental age – are objective measures that are relatively easy to collect in contemporary settings compared to more intensive assessments like parental engagement metrics. This suggests the possibility of developing streamlined screening protocols using readily available data points, which could enhance feasibility in diverse healthcare and educational contexts.

The superior performance of parsimonious models combining selected traditional risk factors with linguistic features suggests the possibility of developing streamlined screening protocols that maintain high predictive accuracy while minimizing administrative burden. However, the implementation of essay-based assessments in contemporary settings requires careful consideration of several factors. The future-oriented nature of our essay prompt (“Imagine that you are now 25 years old…”) may have elicited particularly revealing content about children’s psychological outlook and self-perception. Alternative essay topics or formats, including existing school assignments, might yield different linguistic patterns and predictive value. Additionally, the optimal setting for such assessments—whether integrated into educational curricula, administered during healthcare visits, or implemented through digital platforms—requires further research.

These findings also highlight the potential value of incorporating routine writing exercises in educational settings as a non-invasive means of monitoring children’s psychological well-being, though careful consideration must be given to ethical implications and implementation strategies. While our analysis identified paternal engagement as predictive in the 1960s data, contemporary research by Scott and colleagues confirms that responsive, boundary-setting parenting remains crucial for children’s mental wellbeing despite societal changes(50). This reinforces the continued relevance of integrating family-centered approaches within assessment and intervention frameworks..

### Strength and limitations

This study presents several strengths in its methodological approach and analytical framework. First, the utilization of prospective birth cohort data from the NCDS provides a robust foundation for investigating developmental trajectories, minimizing recall bias and allowing for temporal sequence establishment between predictors and outcomes. Second, our integrated machine learning approach, combining traditional risk factors with natural language processing of children’s essays, represents a novel methodological advancement in pediatric mental health screening. Third, the inclusion of two validated mental health assessment tools (BSAG and Rutter A Scale) strengthens the reliability of our findings through complementary outcome measures. Fourth, our comprehensive consideration of socioeconomic, environmental, and adverse childhood experiences provides a holistic framework for understanding childhood mental health determinants.

However, several limitations warrant consideration when interpreting these findings. First, while the NCDS cohort provides rich historical data, its composition (predominantly white British children born in 1958) may limit generalizability to contemporary, more diverse populations. Second, the mental health measures are constrained by their time period. The Rutter scales were later updated into the Strengths and Difficulties Questionnaire and included items like nail biting, then considered indicative of emotional disturbance but no longer viewed as such(51). Third, some potentially relevant factors such as genetic predisposition, cultural influences, social networks, disability status, and general health conditions were not comprehensively incorporated(10, 11, 52). Fourth, binary classification of mental health outcomes based on threshold scores, while pragmatic for screening, may not fully capture the nuanced spectrum of childhood mental health experiences. Fifth, our machine learning models require validation in contemporary and diverse populations before widespread clinical implementation. Sixth, we excluded children with substantial missing data or very short essays, potentially introducing selection bias. As shown in **Supplementary Table 1**, excluded children demonstrated significantly poorer mental health outcomes and differed on key predictors. This exclusion may have affected model performance, though our integrated approach still demonstrated strong predictive capability (ROC: 0.77-0.81), suggesting potentially greater utility in more diverse populations with pronounced mental health needs. Seventh, we employed internal validation rather than external validation, with feature selection and model optimization performed within the same cohort. While we maintained a separate test dataset, validation on entirely independent cohorts would be more rigorous. This necessitates caution when interpreting performance metrics, as they may not fully generalize to new populations. Eighth, our natural language processing approach treated essays holistically rather than extracting specific linguistic features. While embedding-based approaches capture semantic meaning, their “black box” nature limits interpretation of which writing aspects most strongly predicted mental health outcomes, restricting understanding of underlying psychological mechanisms. Ninth, we focused exclusively on children completing the essay task, potentially introducing selection bias, as children producing very short essays or not completing assignments might systematically differ in mental health profiles. Finally, we did not explore how predictive value of children’s writing might vary across different ages, developmental stages, or essay topics. The future-oriented prompt may have uniquely elicited certain psychological themes, and different writing assignments might yield varying predictive patterns.

## Conclusion

This prospective birth cohort study demonstrates that integrating machine learning with natural language processing of children’s written expressions obviously enhances early detection of potential mental health disorders in school-age children. The optimal predictive performance achieved through combining key environmental predictors with linguistic features suggests a promising pathway for developing efficient screening protocols. While this integrated approach shows potential as a screening tool complementary to clinical assessment, validation in contemporary, diverse populations remains essential. Further research is needed to determine the optimal implementation of essay-based screening across different age groups, cultural contexts, and writing topics before clinical application can be recommended. Our findings offer a practical direction for addressing the diagnostic gap in pediatric mental health care through more accessible and efficient screening methods, potentially enabling earlier intervention for children at risk of mental health disorders.

## Contributors

SC contributed to the concept and study design. SC conducted the analysis. SC, TD, MQ, HL, QAL, AMW, RNC, and TJF. SC drafted the manuscript. TD, MQ, HL, QAL, AMW, RNC, and TJF made critical revisions to the manuscript for important intellectual content. All authors edited and approved the final manuscript.

## Funding

SC’s research was supported by the PENDA, funded by the UK Foreign, Commonwealth and Development Office. RNC’s research was supported by the Medical Research Council (MR/Z504816/1). All research at the Department of Psychiatry in the University of Cambridge is supported by the NIHR Cambridge Biomedical Research Centre (NIHR203312) and NIHR Applied Research Collaboration East of England.

## Role of the funding source

The funder of the study had no role in study design, data collection, data analysis, data interpretation, or writing of the article. The views expressed are those of the author(s) and not necessarily those of the NIHR or the Department of Health and Social Care. For the purpose of open access, the authors have applied a Creative Commons Attribution (CC BY) licence to any Author Accepted Manuscript version arising from this submission.

## Conflict of Interest

RNC consults for Campden Instruments Ltd; receives royalties from Cambridge University Press, Cambridge Enterprise, and Routledge; and is an unpaid non-executive director of Cambridge University Health Partners. TJF’s research group receives funding for methodology consulting from Place2Be, a third sector organisation that provides mental health training and interventions to UK schools. SC and other authors declare no conflict of interest with this work.

## Ethics Statement

This study uses data from the National Child Development Study (NCDS), a nationally representative longitudinal cohort initiated in 1958. Informed consent was obtained for all data collections, with parental consent provided for assessments conducted during childhood, including the age-11 survey and essay-writing task. Formal ethical approval for NCDS follow-ups has been obtained from the UK NHS Multi-Centre Research Ethics Committee (MREC) for all surveys conducted since 2000. Earlier waves, including those in 1958, 1965, 1969, 1974, 1981, and 1991, were conducted prior to the establishment of formal ethics committees or the MREC system. Available documentation indicates that internal ethical review processes were in place for these early waves. For example, the Biomedical Survey and age-55 follow-up were approved by the NHS London-Central Research Ethics Committee (REC) in 2012 (Ref: 12/LO/2010). The current study is a secondary analysis of de-identified public data accessed via the UK Data Service (Study No. 5790). As such, it is exempt from further institutional ethical review under the policies of the London School of Hygiene & Tropical Medicine, as it does not involve human participants or identifiable private information.

## Data Availability Statement

The data that support the findings of this study are openly available in the UK Data Service at http://doi.org/10.5255/UKDA-SN-5790-2, reference number SN: 5790.

## Notes

### Competing Interest Statement

The authors have declared no competing interest.

### Author Declarations

The study used ONLY openly available human data that were originally located at http://doi.org/10.5255/UKDA-SN-5790-2.

