## Supplementary material for "Machine learning and natural language processing for the early detection of potential mental disorders among school-age children: a prospective birth cohort study": Tables and figures

**Figure 1. Participant flow chart.**

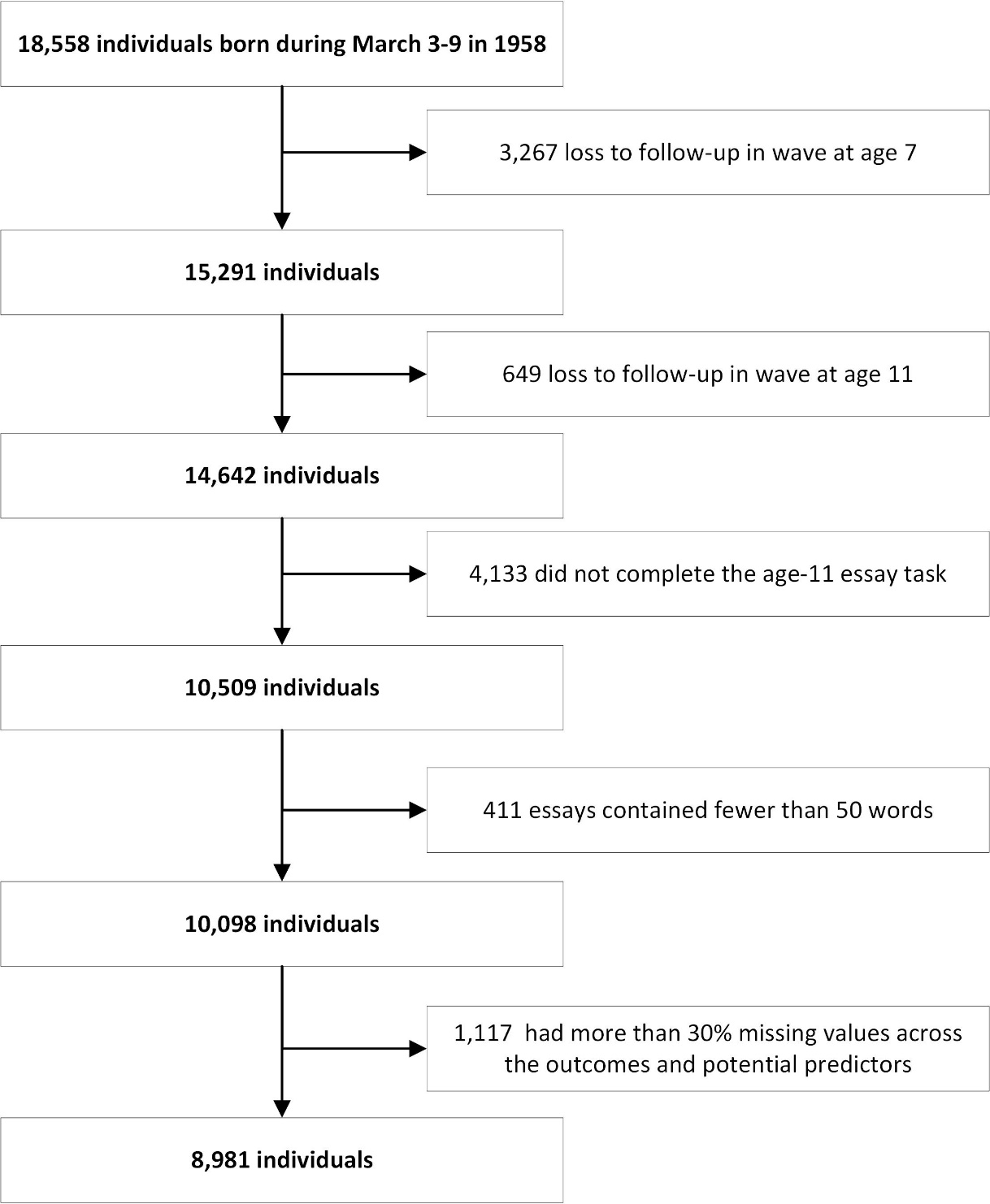

**Table 1**. **Basic description.** Categorical variables were reported as numbers (percentages%) and continuous variables as means (standard deviations).

| **Variables** | **N = 8,981** |
| --- | --- |
| **Socioeconomic position** |  |
| Paternal occupation category at birth (= manual) | 6459 (71.9%) |
| Paternal occupation category at age 7 (= manual) | 6052 (67.4%) |
| Paternal occupation category at age 11 (= manual) | 5793 (64.5%) |
| Father unemployed at age 7 (= yes) | 196 (2.2%) |
| Father unemployed at age 11 (= yes) | 279 (3.1%) |
| Mother education status (= Remained after the compulsory period) | 3200 (35.6%) |
| Father education status (= Remained after the compulsory period) | 2910 (32.4%) |
| Housing tenure at age 7 (= Owned/mortgaged) | 3780 (42.1%) |
| Housing tenure at age 11 (= Owned/mortgaged) | 4135 (46.0%) |
| Number of rooms in the residence at age 7 | 4.78 (1.33) |
| Number of rooms in the residence at age 11 | 4.92 (1.32) |
| Having financial hardships at age 7 (= yes) | 740 (8.2%) |
| Having financial hardships at age 11 (= yes) | 1052 (11.7%) |
| Reported housing difficulties at age 7 (= yes) | 662 (7.4%) |
| Provision of free school meals at age 11 (=yes) | 917 (10.2%) |
| **Adverse childhood experiences** |  |
| Parental separation or divorce (=yes) | 367 (4.1%) |
| Substance misuse by parents (=yes) | 130 (1.4%) |
| Presence of family conflict (=yes) | 564 (6.3%) |
| Parental death (=yes) | 150 (1.7%) |
| Mental health problems among parents (=yes) | 558 (6.2%) |
| Physical neglect experienced from parents (=yes) | 567 (6.3%) |
| Parental involvement in criminal activities (=yes) | 348 (3.9%) |
| **Environmental factors** |  |
| Sex (= female) | 4429 (49.3%) |
| Having diseases at birth (= yes) | 242 (2.7%) |
| Weight category at birth |  |
| Under -2 SD | 199 (2.2%) |
| -2 to -1 SD | 1191 (13.3%) |
| -1 to +1 SD | 6327 (70.4%) |
| +1 to +2 SD | 1020 (11.4%) |
| Over +2 SD | 244 (2.7%) |
| ZBMI at age 7 |  |
| Under -2 SD | 182 (2.0%) |
| -2 to -1 SD | 691 (7.7%) |
| -1 to +1 SD | 6606 (73.6%) |
| +1 to +2 SD | 1141 (12.7%) |
| Over +2 SD | 361 (4.0%) |
| ZBMI at age 11 |  |
| Under -2 SD | 203 (2.3%) |
| -2 to -1 SD | 1169 (13.0%) |
| -1 to +1 SD | 6147 (68.4%) |
| +1 to +2 SD | 1104 (12.3%) |
| Over +2 SD | 358 (4.0%) |
| Mother’s age at child’s birth | 27.58 (5.68) |
| Length of gestational period | 280.74 (12.91) |
| Mother’s parity | 1.30 (1.52) |
| Maternal health conditions during pregnancy (= yes) | 2093 (23.3%) |
| Mother’s marital status at child’s birth (= Married/ Twice married/Cohabiting/Stable Union) | 8693 (96.8%) |
| Mother’s weekly smoking frequency during pregnancy |  |
| None | 6134 (68.3%) |
| 1-9 | 1634 (18.2%) |
| >= 10 | 1213 (13.5%) |
| Mother interested in child education at age 7 (= yes) | 7694 (85.7%) |
| Mother interested in child education at age 11 (= yes) | 7739 (86.2%) |
| Mother engaged in outdoor activities with child at age 7 (= yes) | 8840 (98.4%) |
| Mother engaged in outdoor activities with child at age 11 (= yes) | 8370 (93.2%) |
| Mother participated in reading activities with child at age 7 (=yes) | 7535 (83.9%) |
| Father’s age at child birth | 30.65 (6.36) |
| Father interested in child education at age 7 (= yes) | 6998 (77.9%) |
| Father interested in child education at age 11 (= yes) | 6953 (77.4%) |
| Father engaged in outdoor activities with child at age 7 (= yes) | 8431 (93.9%) |
| Father engaged in outdoor activities with child at age 11 (= yes) | 7916 (88.1%) |
| Father participated in reading activities with child at age 7 (= yes) | 6443 (71.7%) |
| Father participated in childcare at age 7 (=yes) | 7960 (88.6%) |
| Father participated in childcare at age 11 (=yes) | 8001 (89.1%) |
| **Outcomes** |  |
| Scores of Bristol Social Adjustment Guide (BSAG) at age 11 | 15.81 (17.01) |
| Scores of Rutter A Scale at age 11 | 6.56 (3.65) |

**Figure 2. Relative importance of predictors for childhood mental health risk.** Data presents the relative importance of selected predictor variables in forecasting potential mental health disorders among school-aged children, based on the Bristol Social Adjustment Guide (BSAG) score at age 11. The x-axis displays the importance score, representing each variable's contribution to the model's predictive accuracy, while the y-axis lists the predictor variables in descending order of importance. The importance scores were derived using recursive feature elimination with cross-validation in the training dataset. Higher scores indicate greater predictive value for identifying potential mental health disorders.

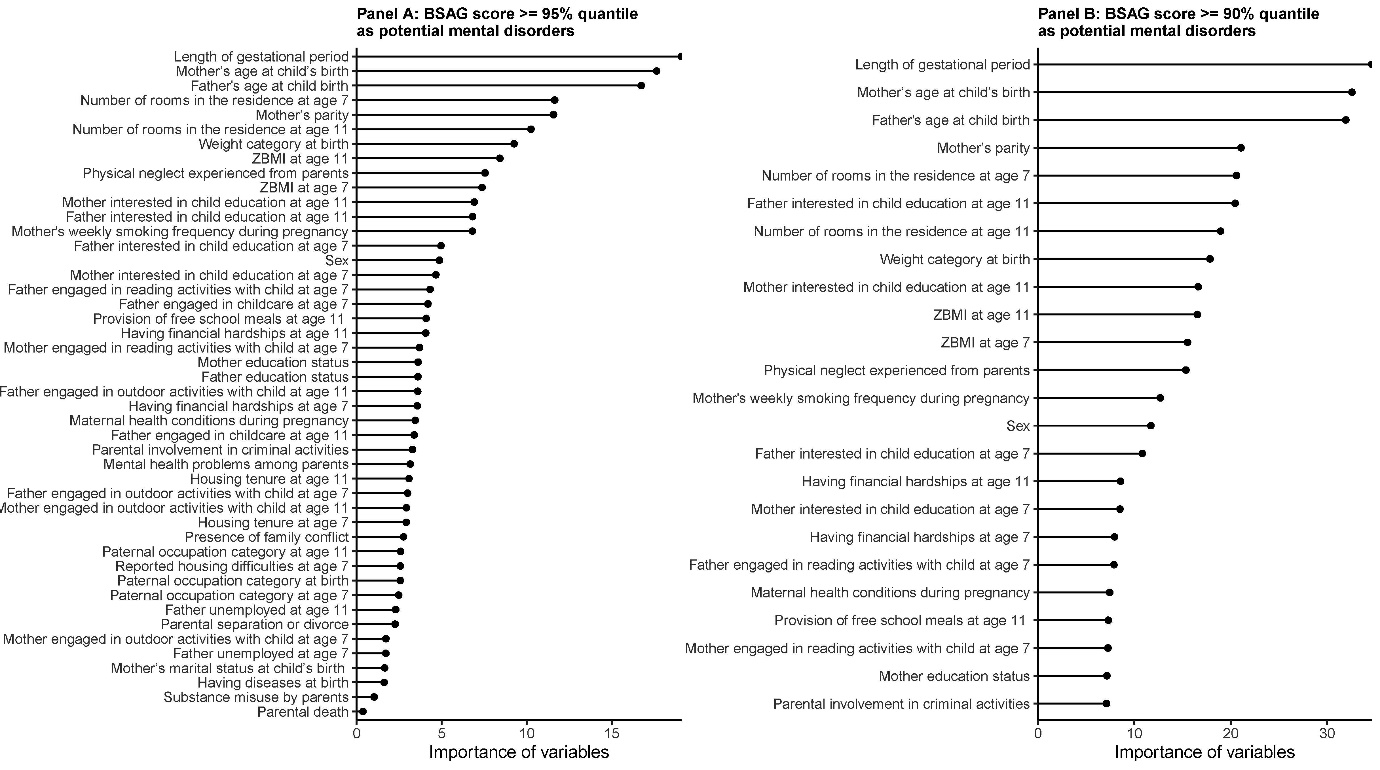

**Figure 3. Receiver operating characteristic (ROC) curves comparing predictive performance of machine learning models for childhood mental health assessment.** The figure presents ROC curves demonstrating the predictive performance of six different modeling approaches: stand-alone models using either all selected variables or essay features, and combined models integrating essay features with varying numbers of top selected variables (5, 10, 15, and 20). The x-axis shows the false positive rate (1 - specificity), and the y-axis displays the true positive rate (sensitivity). Each curve represents the best-performing model within its respective modeling approach, as determined by the highest ROC value.

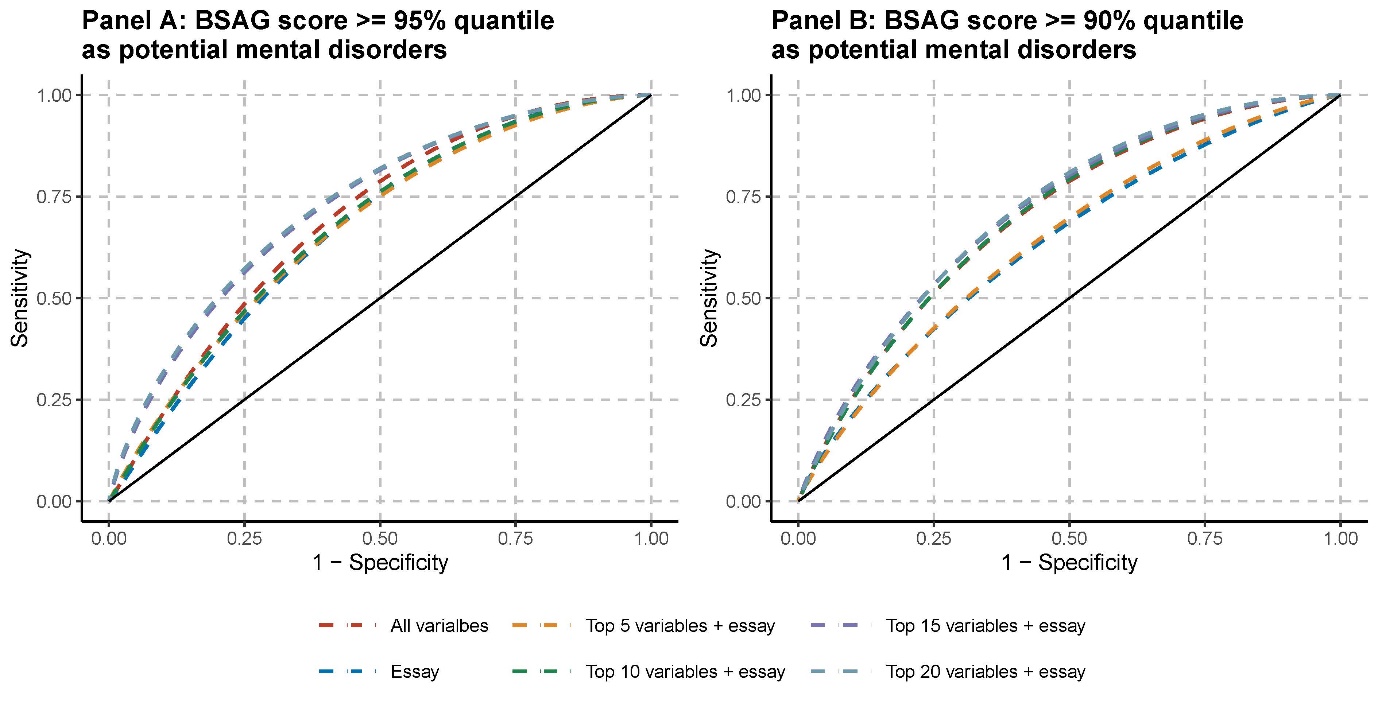

**Table 2. Performance metrics of machine learning models using different predictor combinations for mental health risk detection.** Data summarizes the performance metrics of machine learning models predicting potential mental health disorders based on Bristol Social Adjustment Guide (BSAG) scores at age 11, using two different thresholds (95th and 90th percentiles). The models were evaluated using different combinations of predictors: traditional variables alone, essay features alone, and varying combinations of top selected variables (5, 10, 15, or 20) with essay features. For each predictor combination shown, we selected only the best-performing model out of eight different algorithms tested (linear discriminant analysis, logistic regression, XGBoost, k-nearest neighbors, neural networks, naive Bayes, support vector machines, and random forest). The best model was determined by the highest ROC value in the test dataset.

| **Outcomes** | **predictors** | **ROC** | **Sensitivity** | **Specificity** | **Accuracy** | **AUC** | **F** | **Kappa** | **Precision** | **Recall** |
| --- | --- | --- | --- | --- | --- | --- | --- | --- | --- | --- |
| BSAG scores >= 95% quantile as potential mental disorders | all selected variables | 0.70 (0.63, 0.76) | 0.56 (0.45, 0.67) | 0.74 (0.72, 0.76) | 0.73 (0.71, 0.75) | 0.12 (0.06, 0.17) | 0.16 (0.12, 0.20) | 0.09 (0.05, 0.12) | 0.09 (0.06, 0.12) | 0.56 (0.45, 0.67) |
|  | essay | 0.67 (0.60, 0.74) | 0.63 (0.53, 0.74) | 0.62 (0.60, 0.65) | 0.62 (0.60, 0.65) | 0.10 (0.06, 0.14) | 0.13 (0.10, 0.16) | 0.05 (0.03, 0.08) | 0.07 (0.05, 0.09) | 0.63 (0.53, 0.74) |
|  | top 5 selected variables + essay | 0.77 (0.71, 0.83) | 0.66 (0.55, 0.77) | 0.72 (0.70, 0.74) | 0.72 (0.70, 0.74) | 0.24 (0.15, 0.34) | 0.17 (0.13, 0.21) | 0.10 (0.07, 0.14) | 0.10 (0.07, 0.12) | 0.66 (0.55, 0.77) |
|  | top 10 selected variables + essay | 0.77 (0.72, 0.83) | 0.61 (0.50, 0.72) | 0.78 (0.76, 0.80) | 0.77 (0.75, 0.79) | 0.28 (0.19, 0.39) | 0.19 (0.14, 0.23) | 0.12 (0.08, 0.16) | 0.11 (0.08, 0.14) | 0.61 (0.50, 0.72) |
|  | top 15 selected variables + essay | 0.81 (0.76, 0.86) | 0.62 (0.51, 0.73) | 0.78 (0.76, 0.80) | 0.77 (0.75, 0.79) | 0.37 (0.28, 0.49) | 0.19 (0.15, 0.24) | 0.13 (0.09, 0.17) | 0.11 (0.08, 0.14) | 0.62 (0.51, 0.73) |
|  | top 20 selected variables + essay | 0.81 (0.76, 0.86) | 0.63 (0.52, 0.74) | 0.78 (0.76, 0.80) | 0.77 (0.75, 0.79) | 0.38 (0.28, 0.50) | 0.20 (0.15, 0.25) | 0.13 (0.09, 0.18) | 0.12 (0.09, 0.15) | 0.63 (0.52, 0.74) |
| BSAG scores >= 90% quantile as potential mental disorders | all selected variables | 0.70 (0.65, 0.74) | 0.58 (0.51, 0.65) | 0.74 (0.72, 0.76) | 0.73 (0.71, 0.75) | 0.26 (0.20, 0.32) | 0.30 (0.25, 0.34) | 0.17 (0.13, 0.22) | 0.20 (0.16, 0.24) | 0.58 (0.51, 0.65) |
|  | essay | 0.64 (0.59, 0.68) | 0.68 (0.61, 0.75) | 0.54 (0.51, 0.56) | 0.55 (0.53, 0.58) | 0.15 (0.12, 0.19) | 0.23 (0.20, 0.27) | 0.08 (0.05, 0.11) | 0.14 (0.12, 0.16) | 0.68 (0.61, 0.75) |
|  | top 5 selected variables + essay | 0.81 (0.78, 0.85) | 0.74 (0.68, 0.81) | 0.73 (0.71, 0.75) | 0.73 (0.71, 0.75) | 0.39 (0.32, 0.47) | 0.35 (0.31, 0.40) | 0.24 (0.20, 0.28) | 0.23 (0.20, 0.27) | 0.74 (0.68, 0.81) |
|  | top 10 selected variables + essay | 0.82 (0.79, 0.85) | 0.64 (0.57, 0.71) | 0.78 (0.76, 0.80) | 0.76 (0.74, 0.78) | 0.48 (0.42, 0.56) | 0.35 (0.30, 0.40) | 0.24 (0.19, 0.29) | 0.24 (0.20, 0.28) | 0.64 (0.57, 0.71) |
|  | top 15 selected variables + essay | 0.82 (0.78, 0.85) | 0.70 (0.64, 0.77) | 0.78 (0.76, 0.80) | 0.77 (0.75, 0.79) | 0.46 (0.39, 0.53) | 0.38 (0.33, 0.43) | 0.27 (0.22, 0.32) | 0.26 (0.22, 0.30) | 0.70 (0.64, 0.77) |
|  | top 20 selected variables + essay | 0.82 (0.79, 0.85) | 0.72 (0.65, 0.79) | 0.77 (0.75, 0.79) | 0.77 (0.75, 0.79) | 0.46 (0.39, 0.54) | 0.38 (0.33, 0.43) | 0.28 (0.23, 0.33) | 0.26 (0.22, 0.30) | 0.72 (0.65, 0.79) |
